# Feasibility and Preliminary Effectiveness of a Discrete Choice-Based Pre-Exposure Prophylaxis Delivery Model for Female Sex Workers in Uganda

**DOI:** 10.1101/2025.11.20.25340720

**Authors:** Ruth Mpirirwe, Andrew Mujugira, Bosco B. Agaba, Joan Nangendo, Mbabazi Lenin Dennis, Fred C. Semitala, Peter Kyambadde, Joan Kalyango, Charles Karamagi, Agnes Kiragga, Anne R. Katahoire, Moses R Kamya

## Abstract

The current PrEP delivery models in Uganda do not fully address the socio-economic, cultural, and logistical challenges faced by female sex workers (FSW), resulting in low adherence, mistrust in the healthcare system, and missed opportunities for effective PrEP care, as reflected in national data. This study evaluated the feasibility and effectiveness of a discrete-choice PrEP delivery model to improve PrEP uptake, adherence, and outcomes among FSWs in Kampala, Uganda. We conducted a discrete choice experiment in October–November 2023 and identified an optimal model combining facility-based, health worker-led, in-person support with short message service (SMS) reminders. From March to May 2024, we implemented a pilot study to assess feasibility and preliminary effectiveness. We recruited 49 FSWs using peer-led snowball sampling at the Most At-Risk Population Initiative (MARPI) clinic at Mulago Hospital. We evaluated effectiveness using a pre-post design, comparing adherence scores at one and two months. Feasibility was measured using six 4-point Likert-scale items (score range 6–24; higher scores = greater feasibility). All 50 participants completed the follow-up. The median age was 27 years (IQR 21–30); most were Christian (86%), had primary education (55%), were single (76%), had sex with males (79%), and were in casual relationships (88%). Median adherence improved from 0.90 (IQR 0.83–0.93) at baseline to 1.00 (IQR 0.97–1.00) at two months. All 50 participants returned for their refill at the 2-month visit. Feasibility was high, with a mean score of 21.26 (SD 1.03). Integrating in-person facility-based support with SMS reminders significantly improved oral PrEP adherence among high-risk FSW. Improved adherence, full retention, and strong feasibility demonstrate the promise of combining face-to-face health care with mobile technology. Future research should examine the long-term effectiveness and scalability of this discrete-choice-informed model to sustain PrEP adherence among FSW.

## INTRODUCTION

In Uganda, 77% of female sex workers (FSWs) initiate HIV pre-exposure prophylaxis (PrEP) when offered.^1^ However, persistence remains a challenge, as only 19% of FSWs return for refill appointments.^2^ This significant decline in sustained prevention results form the unique, complex challenges that hinder continuous oral PrEP use among FSWs, including the burden of daily dosing, confusion of PrEP with HIV treatment, anticipated side effects, and multi-level stigma.^3–7^ Although high initiation targets are being achieved and FSWs receive strong policy emphasis (40% of PrEP initiators in Uganda are FSWs), PrEP program optimization remains essential, as this initial success masks health system fragility in sustained engagement. National programme priorities should therefore shift from maximizing initiation numbers to ensuring high adherence and long-term retention.

The current PrEP delivery models in Uganda do not sufficiently address the distinct socio-economic, cultural, and logistical challenges faced by FSW. Current approaches typically use a one-size-fits-all strategy, which inadequately addresses FSW concerns related to privacy and confidentiality, fear of judgment from healthcare providers and peers, time constraints due to nocturnal work schedules, mobility challenges that hinder attendance for scheduled visits, and the absence of regular follow-ups essential for maintaining high adherence rates.^8,9^ As a result, these factors contribute to low adherence, mistrust in the healthcare system, and missed opportunities for adequate care, as reflected in the national PrEP data. Involving FSW in the development of PrEP delivery models could enhance retention in care. However, to date, no discrete choice experiments (DCEs)^10^ have been conducted in Uganda involving FSWs to better comprehend their specific preferences regarding PrEP delivery.

The Discrete Choice Experiment is a research method used to evaluate the trade-offs that individuals are willing to make between different attributes of an intervention by simulating real-world decision-making scenarios^11^. In this study, we used the DCE approach to obtain data on the attributes of a PrEP delivery model that influence FSW decisions to access and adhere to PrEP in Uganda. The attributes examined in the DCE included types of healthcare service delivery, location, and adherence support. By applying DCE to PrEP delivery, we aimed to better align with FSWs’ preferences and needs of PrEP delivery in Uganda^12^. This study sought to evaluate the feasibility of this tailored PrEP delivery model in improving PrEP uptake, adherence, and effectiveness among FSWs in Kampala, Uganda.

## METHODS

### Study Population and Setting

We previously used a discrete-choice experiment to identify an optimal PrEP delivery model (i.e., facility-based, health worker-led, or in-person with short message service [SMS] reminders)^13^ at the Most At-Risk Population Initiative (MARPI) clinic within the Mulago National Referral Hospital complex in Kampala, Uganda, from March to May 2024. We recruited 50 FSWs from the MARPI clinic, which serves an estimated 10,000 FSWs annually, into a pilot study to evaluate the feasibility and preliminary effectiveness of this PrEP delivery model. Before participating in the study, FSWs received information about the study’s objectives and procedures. Using a pre-posttest design, we assessed participants’ adherence scores before and after the intervention to determine whether the facility/health worker/in-person with short message service (SMS) reminder model influenced changes in oral PrEP adherence.^14^ This approach was preferred due to its feasibility and suitability for evaluating the model’s preliminary effectiveness, particularly given the small participant size. It enabled the identification of implementation changes that needed to be addressed before scaling up the intervention. The pre–post design was also appropriate as it efficiently provided insights into whether the intervention was feasible, acceptable, and potentially beneficial, while laying the groundwork for more rigorous studies.

### Inclusion Criteria

Participants were FSWs engaged in transactional sex, who met the following criteria: (1) provided informed consent to participate in the study; (2) were aged 14 years or older, including emancipated or mature minors; (3) committed to participating for the two-month study period; (4) were willing to complete study procedures; (5) were available during the data collection period; (6) were MARPI clients; (7) spoke and understood English or Luganda; and (8) were HIV-negative.

### Exclusion Criteria

In this study, FSWs who were participating in another PrEP or HIV prevention study, were very ill, allergic to TDF, 3TC, FTC, or any other PrEP drug, had hepatitis B virus infection (self-report or medical records), or had chronic kidney disease (self-report or medical records), were excluded.

### Population and procedures

All FSWs who met the inclusion criteria were sequentially selected until the sample size of 50 was reached. The PrEP candidates were offered and initiated on PrEP by MARPI PrEP counselors at either the clinic or community distribution points. We anticipated that at least 20% of all FSW attending the clinic would be eligible for PrEP, and at least 30% would initiate it^15,16^. We predicted that some who started PrEP would continue taking it for the 2-month study period.

#### PrEP Initiation

HIV-negative FSWs who were interested and eligible for PrEP were initiated on PrEP on the same day. Before initiation, PrEP counsellors provided risk-reduction counselling and PrEP medication-adherence counselling according to national PrEP guidelines. Oral PrEP was dispensed by PrEP counselors. In Uganda, oral PrEP is taken as a daily pill and is available as a 30-day pack of either TDF/FTC 300/200mg (preferred option) or TDF/3TC 300/300mg (alternative).^15^ The study site provided PrEP.

#### PrEP retention

Follow-up occurred throughout the 2-month study period, in accordance with national PrEP guidelines.^17^ All study participants were initially assigned to the conventional delivery models in the first month. In contrast, during the second month, FSWs were introduced to the DCE-informed PrEP delivery model, i.e, facility/health worker/in-person with SMS reminders. Data collection over the two months was based on adherence support outlined in the PrEP technical guidelines. None of the FSW acquired HIV during the study period. Strategies for maintaining enrolled FSWs on PrEP included sending SMS/text reminders to FSWs.

#### PrEP discontinuation

None of the FSW discontinued PrEP during the study period. The majority of the respondents resided within the Central Region. The completed questionnaires were assessed for completeness by the Principal Investigator.

#### Data collection

The Ministry of Health (MoH) PrEP facility record was used to collect data on age, religion, education level, marital status, sexual orientation, and type of sexual partner. Data was collected for two months by two PrEP providers at the study site, assisted by one research assistant. The study team received training on the study protocol, data collection tools, and study procedures before data collection.

#### Quality Control

Pre-testing of the data collection tools was conducted on a sample of approximately 20 participants to evaluate the appropriateness of the variables. The research assistants, who held Master’s degrees, double-entered the data to minimise analysis errors. We translated the feasibility questions into Luganda, given that most FSW come from the central region.

### Statistical Analysis

The primary outcomes were the feasibility and effectiveness of the DCE-informed model. The feasibility of the facility/healthworker/in-person with SMS reminders PrEP delivery model. We assessed feasibility using a set of 6 4-point Likert-scale questions, scored 1-4, with higher scores indicating greater feasibility. We defined feasibility as a score above the median (15) of the possible scores (6-24). Pill count was used as a measure of oral PrEP adherence to assess the effectiveness of this PrEP delivery model. We compared the adherence scores (i.e., pills dispensed-pill returned)/days between visits) before and after the intervention using the Mann-Whitney U test. We measured retention as the proportion of enrolled FSWs who attended the 2-month follow-up visit after the implementation of this PrEP delivery model.

### Ethics Approval

We obtained approval for the study from the Makerere University School of Medicine Research Ethics Committee (Mak-SOMREC-2022-299) and the Uganda National Council for Science and Technology (SS1223ES). We obtained administrative from Makerere University’s Clinical Epidemiology Unit and Mulago National Referral Hospital Ethics Committee. All participants provided written informed consent and received an IRB-approved reimbursement of 20,000 Uganda Shillings ($5.30) for their time, effort, and transportation costs.

## RESULTS

### Characteristics of study participants

The median age of study participants was 27 years (interquartile range [IQR] 21-30). Most FSWs (86%, 42/49) identified as Christian, and 56% (27/48) had completed primary education, i.e., seven years of schooling. A majority (88% 43/49) reported having casual sexual partners (**Table 1**). Retention in PrEP care was 100% at 2 months into the intervention.

**Table 1:** Baseline Characteristics of FSW (N=49)

| Variable and categories | Frequency (N, %) |
| --- | --- |
| **Age (years)** | 27 (21, 30) |
| **Religion\*** |  |
| Christian | 42 (85.7) |
| Moslem | 7(14.3) |
| **Education level<sup>#</sup>** |  |
| None | 2(4.1) |
| Primary level | 27(55.1) |
| Secondary level | 19(38.8) |
| Tertiary level | 1(2.0) |
| **Marital status** |  |
| Cohabiting | 1(2.0) |
| Divorced | 1(2.0) |
| Married | 7(14.0) |
| Separated | 4(8.0) |
| Single | 36(72.0) |
| Widowed | 1(2.0) |
| **Had sex with<sup>#</sup>** |  |
| Females | 10(20.8) |
| Males | 38(79.2) |
| **Type of sexual relationship\*** |  |
| Casual partners | 43(87.8) |
| Regular partners | 1(2.0) |
| Both casual and regular | 5(10.2) |

\*N=49, #N=48

### Preliminary effectiveness of DCE-informed PrEP delivery model assessed through adherence and retention

The implementation of the preferred PrEP delivery model significantly increased oral PrEP scores from 0.90 (IQR 0.83, 0.93) to 1.00 (0.97, 1.00) (p<0.001). We observed similar significant increases in adherence to oral PrEP across subgroups of FSW, including adolescent girls and young women (AGYW) (p<0.001), women older than 24 years (p<0.001), and by religion and sex orientation (p<0.001). However, we observed no significant change in those with no formal education (p=0.18) or in those with a tertiary education level (p-value=0.32). Participants with a regular partner had no change in adherence scores (P value 0.32) **(Table 2)**.

**Table 2:** PrEP adherence scores pre- and post-intervention.

| **Variable with categories** | **Baseline adherence score**<br>**Median (IQR)** | **Post-intervention adherence score**<br>**Median (IQR)** | **P-value** |
| --- | --- | --- | --- |
| Overall adherence | 0.90 (0.83, 0.93) | 1.00 (0.97, 1.00) | <0.001 |
| **Sub-groups of FSW** |  |  |  |
| AGYW* (15-24 years) | 0.89 (0.83, 0.93) | 0.99 (0.97, 1.00) | <0.001 |
| Older FSW (>24 years) | 0.90 (0.83, 0.92) | 1.00 (0.97, 1.00) | <0.001 |
| **Religion** |  |  |  |
| Christian | 0.87 (0.83, 0.93) | 1.00 (0.97, 1.00) | <0.001 |
| Moslem | 0.90 (0.90, 0.93) | 1.00 (1.00, 1.00) | 0.02 |
| **Years of Schooling** |  |  |  |
| None | 0.92 (0.87, 0.97) | 1.00 (1.00, 1.00) | 0.18 |
| Primary level (0-7) | 0.90 (0.83, 0.93) | 1.00 (0.97, 1.00) | <0.001 |
| Secondary level (8-13) | 0.90 (0.87, 0.93) | 1.00 (0.97, 1.00) | <0.001 |
| Tertiary level (>13) | 0.87 (0.87, 0.87) | 1.00 (1.00, 1.00) | 0.32 |
| **Client's sex** |  |  |  |
| Female | 0.87 (0.83, 0.97) | 0.99 (0.97, 1.00) | 0.005 |
| Male | 0.90 (0.87, 0.93) | 1.00 (0.97, 1.00) | <0.001 |
| **Partner type** |  |  |  |
| Casual partners | 0.87 (0.83, 0.90) | 1.00 (0.97, 1.00) | <0.001 |
| Regular partners | 0.97 (0.97, 0.97) | 1.00 (1.00, 1.00) | 0.32 |
| Both casual & regular | 0.93 (0.93, 0.93) | 1.00 (1.00, 1.00) | 0.042 |

### Feasibility of DCE-informed PrEP delivery

On average, participants rated feasibility at 21.26 out of 30 on the feasibility scale; scores ≥15/24 were considered high. The responses were tightly clustered around the mean (SD 1.03), indicating that most participants’ scores were very close to 21.26, suggesting consistency in their perception of feasibility **(Table 3).**

**Table 3:** Participants’ rating of preferred PrEP delivery model.

| Construct of feasibility | Mean (SD) | Reference category |
| --- | --- | --- |
| Overall feasibility | 21.26 (1.03) | High |
| Was not burdened by the PrEP delivery model | 3.32 (0.51) | High |
| Would prefer to continue with this PrEP delivery model | 3.60 (0.50) | High |
| Belief that nurses can effectively deliver PrEP services | 3.50 (0.58) | High |
| This PrEP model was more convenient than others | 3.50 (0.58) | High |
| This model was not costly from the users' perspective | 3.56 (0.50) | High |

## DISCUSSION

This discrete-choice experiment involving 49 FSW in Uganda suggests that combining in-person care from facility-based nurses with text message reminders improved oral PrEP adherence. The intervention was effective across different religious groups and among clients of different sexes, indicating its versatility and potential for widespread implementation among diverse groups of FSWs. The combination of facility-based care and mobile health technology (SMS reminders) offers a practical, scalable solution for improving PrEP adherence in real-world settings. This notable improvement in adherence was observed across several key subgroups, indicating the model’s broad applicability. Specifically, both AGYW and older FSW showed remarkable increases in adherence.

Decentralized and tailored care helps address the structural barriers faced by FSWs, meeting their needs for convenience, confidentiality, and stigma-free care. Our findings align with prior research that highlights the importance of tailored interventions for high-risk populations^18^. The addition of regular SMS reminders, alongside in-person facility-based care, appeared to address the barriers to access and continuity of care that FSWs often face. Regular text reminders serve as behavioral cues that mitigate forgetfulness and help normalize PrEP use amid competing daily priorities, stigma, and mobility common in this population. A scoping review evaluated mHealth interventions supporting PrEP use among adolescent girls and young women in sub-Saharan Africa, identifying SMS reminders, mobile applications, telecounselling, and web-based platforms as the most common tools, generally acceptable and feasible, for discreet support, reminders, and linkage to PrEP services. The review underscores the need for well-designed implementation studies that measure real-world engagement and effectiveness, and tailor digital support within youth-friendly service models.^19^ Coupled with the facility-based backing where trained providers offer personalized counselling, refills, and laboratory follow-up, this delivery model created a reinforcing loop of motivation, accountability, and social connection to care. This dual approach aligns with evidence that multimodal adherence support is more effective than single-component interventions in sustaining medication use among key populations. Similar results found that SMS reminders improved adherence and appointment keeping among PrEP users, while the non-judgmental facility-based services were critical to retention among FSWs.^20–22^

Consistent with other studies, our model’s effectiveness was observed across different religious groups and regardless of the clients’ sex.^23–25^ This broad applicability suggests that the intervention effect did not vary significantly due to these demographic factors, thereby supporting the feasibility of implementing this model across diverse populations. Similarly, the study revealed essential findings regarding specific subgroups in which the intervention had a limited impact. For example, no significant change was observed in adherence scores for participants with no formal education and those with tertiary education. This suggests that other underlying variables, such as socio-cultural contexts or individual perceptions of healthcare, may have a more profound influence on adherence behaviors, extending beyond formal educational attainment.^23^ Recent evidence from sub-Saharan Africa suggests the relationship between education and PrEP adherence is inconsistent. Among Ugandan FSWs, a 2025 mixed-methods study found no independent association between education level and continuation (a proxy for sustained adherence) after adjustment, underscoring the primacy of structural and service-delivery factors over schooling alone^24^. In contrast, a study found that higher education is consistently associated with better PrEP knowledge and more favorable attitudes, which may facilitate, but do not guarantee, adherence. For instance, social support networks and logistical considerations have been identified as critical factors impacting adherence trajectories, particularly among vulnerable populations such as AGYW.^25^ Understanding the multifaceted interplay between internal drivers and external circumstances, such as PrEP delivery models that strengthen individual motivation and self-efficacy, enhance social support networks, and address structural barriers like stigma, access, and affordability. For instance, adherence programs could combine counselling and behavioral support (targeting internal motivation) with mobile health reminders, peer-led follow-up, and differentiated service delivery models (addressing external facilitators and constraints).

This integrated approach ensures that both personal and contextual determinants of adherence are holistically addressed, which is a crucial step in developing effective PrEP adherence strategies.^26–29^ Interestingly, participants with a regular partner also showed no significant change in adherence scores. This finding suggests that relationship dynamics, including regular partnership status, did not directly influence adherence to oral PrEP in this pilot. In contrast, evidence from multiple studies indicates that partner support, communication, trust, and the nature of sexual arrangements are critical determinants of PrEP adherence outcomes.^30,31^ For instance, among Ugandan HIV serodifferent couples, intimate partner violence (IPV) was negatively associated with PrEP adherence, underscoring how maladaptive relationship dynamics can undermine prevention efforts.^32^ Similarly, in African women with HIV Sero different relationships, those reporting economic or physical IPV had significantly lower PrEP adherence levels compared to those not experiencing abuse.^33^ Conversely, supportive partnerships facilitate better adherence. In Malawi, male partners’ active involvement, including shared decision-making and emotional encouragement, was positively linked to pregnant women’s PrEP uptake and continued use.^34^ This aligns with findings from Kenya and Uganda^35^, where disclosure of oral PrEP use to partners and perceived partner approval improved adherence behaviors.

Perceived stigma within relationships also affects adherence. Some individuals avoid initiating or continuing PrEP due to fears that their partners may interpret its use as a sign of infidelity or promiscuity. It is possible that, for individuals in stable relationships, the perceived need for PrEP could be lower, or that other relationship-related factors, such as support or opposition, communication quality, trust, IPV, and stigma, may be impacting adherence. This observation warrants further investigation into the interplay between relationship context and PrEP adherence^36–38^. Overall, this study underscores the importance of combining facility-based care with mobile health interventions, such as SMS reminders, to improve oral PrEP adherence. However, the varying responses across subgroups emphasize the need for further customization of interventions better to address the specific challenges and needs of FSW. More targeted approaches that account for factors such as education level and relationship context may be needed to maximize the impact of PrEP adherence programs. The present study, therefore, adds to a growing body of evidence supporting hybrid models that combine digital and face-to-face support to strengthen PrEP adherence and continuity of use in real-world contexts.

## STRENGTHS AND LIMITATIONS

This study is among the first to apply a discrete–choice–based delivery approach to PrEP in a real-world setting among FSWs in Uganda. This design enabled participants to make informed choices among different service delivery attributes, thereby enhancing acceptability and alignment with end-user preferences. By linking stated preferences from earlier DCE to an actual intervention model, the study bridged the gap between theoretical preference data and practical program implementation, a crucial methodological advancement in HIV prevention research. The intervention targeted FSWs, who remain at disproportionately high risk for HIV infection, generating evidence specific to this population, which provides valuable insights for tailoring PrEP delivery strategies to FSWs in Uganda and similar settings. The model demonstrated strong retention and adherence outcomes over the pilot period, indicating that combining in-person facility support with SMS reminders is operationally feasible and acceptable in resource-limited contexts. The sample size and scope of this pilot study limit the generalizability of the findings. Larger studies across multiple settings and with more diverse populations are needed to confirm these results and ensure broader applicability. The study may have focused primarily on short-term adherence, and it remains unclear whether the intervention will sustain its impact over time. Longitudinal studies are needed to evaluate the long-term effectiveness and sustainability of this intervention.

## CONCLUSIONS AND RECOMMENDATIONS

Integrating in-person support from facility-based nurses with text message reminders significantly improved oral PrEP adherence among this group of high-risk female sex workers. The rise in adherence scores, absence of participant dropout, and the practical implementation across various subgroups of sex workers underscore the promise of merging face-to-face healthcare with mobile technology. Future studies should evaluate the long-term effectiveness of this DCE-informed approach in sustaining oral PrEP adherence within this population.

## Data Availability

The datasets used and analyzed during the current study are available from the corresponding author on reasonable request.

## AUTHORS’ CONTRIBUTIONS

Study conceptualization - RM, JK, CK, AK, ARK, MRK, and AM. Protocol development - RM and BBA. Data collection process and analysis - RM, BBA, MLD, and JN. First manuscript draft - RM and MRK. Manuscript revisions - RM, JN, ARK, MRK, and AM. Supervision of the data collection process and data management (RM). Overall study supervision - ARK, MRK, and AM. All authors read and approved the final manuscript.

## CONSENT FOR PUBLICATION

Not applicable.

